# Symptoms of depression and anxiety among Vietnamese immigrants in Japan during the COVID-19 pandemic: a cross-sectional web-based study

**DOI:** 10.1101/2022.03.09.22271973

**Authors:** Tadashi Yamashita, Pham Nguyen Quy, Emi Nogami, Erina Seto-Suh, Chika Yamada, Saori Iwamoto, Kyoko Shimazawa, Kenji Kato

**Author notes:** Corresponding author: Tadashi Yamashita.

## Abstract

**Background:** Mental health among immigrants in Japan may be related to aspects of COVID-19 infection as well as pandemic-related lifestyle changes, investigating mental health status among Vietnamese residents remains an important public health concern. The mental health status of Vietnamese immigrants in Japan during the COVID-19 epidemic remains unclear. We aimed to examine the mental health status and related factors among Vietnamese immigrants in Japan during the coronavirus disease 2019 (COVID-19) pandemic using data from an online cross-sectional survey administered from September 21 to October 21, 2021.

**Methods:** Patient Health Questionnaire-9 (PHQ-9) and Generalized Anxiety Disorder 7-item (GAD-7) scores were considered the main outcome measures. Multivariable logistic regression was used to identify factors related to symptoms of depression and anxiety, and the chosen variables were entered simultaneously in the survey.

**Results:** Among 621 participants (age: 26.0±4.8 years; male: 347 [55.9%]) who completed the questionnaire, 73.7% reported a decrease in income when compared with the period before the COVID-19 pandemic, and 60.4% reported being recently affected by poor socioeconomic status. Moderate to severe symptoms of depression (PHQ-9 score ≥ 10 points) and mild-to-severe symptoms of anxiety (GAD-7 score ≥5 points) were observed in 203 (32.7%) and 285 (45.9%) individuals, respectively. Factors related to depressive symptoms were age (odds ratio [OR]=0.94, p=0.043), pre-existing health conditions (OR=2.46, p<0.001), and subjective socioeconomic status (OR=2.47, p<0.001). Factors related to anxiety symptoms were being single (OR=1.72, p=0.044), pre-existing health conditions (OR=2.52, p<0.001), subjective socioeconomic status (OR=2.72, p<0.001), and a partner with whom to discuss one’s health (OR=1.66, p=0.013).

**Conclusions:** The current findings demonstrate that, when compared with non-Vietnamese Japanese people, people with Vietnamese backgrounds experienced a decrease in income, worsening working conditions, and poor mental health status during the COVID-19 pandemic. These findings highlight the potential contribution of low socioeconomic status and social isolation to poor mental health status.

## Background

The number of foreign residents in Japan has continued to increase annually, totalling approximately 2,760,000 as of December 2021. Specifically, there has been a rapid increase in the number of Vietnamese immigrants, the population of which has increased from 10,000 in 1990 to 430,000 in 2021. [1] The Immigration Services Agency of Japan recently reported that the country includes the second-largest number of Vietnamese immigrants after China, which can be attributed to the increase of technical interns and skilled labour positions adopted in Japan. [2]

The coronavirus disease 2019 (COVID-19) pandemic has had a severe impact on health and society worldwide. In Japan, approximately 17 million people have been infected with COVID-19, resulting in approximately 36,000 deaths by July 2022. [3] The pandemic has also affected the lives of Vietnamese immigrants living in Japan, who already hold a marginalised and precarious position, by reducing working hours and increasing layoffs. Furthermore, temporary restrictions on returning to Vietnam may have significantly affected mental health among immigrants.

Studies on mental health due to the COVID-19 pandemic showed that people with chronic illnesses and poor job prospects [4] and women suffered more mental distress than men. [5] On the other hand, social capital has been reported to play a protective function in mental health. [6] Studies of immigrants during the COVID-19 pandemic have reported mental health crises among immigrants in the US, [7] Canada, [8] Europe, [9] India, [10] and South Korea. [11] A web-based survey on mental health of immigrants in 2021 reported most of students from China in Japan have worries about being infected regarding COVID-19 prevention, [12] and approximately 70% of Chinese immigrants in Japan have expressed concerns regarding COVID-19 infection. [13] However, the mental health status of Vietnamese immigrants in Japan during the COVID-19 epidemic remains unclear. Vietnamese immigrants in Japan may have similar concerns regarding COVID-19.

Since mental health among immigrants in Japan may be related to aspects of COVID-19 infection as well as pandemic-related lifestyle changes, investigating mental health status among Vietnamese residents—the second-largest foreign community in Japan—remains an important public health concern. Examining the relationship between socioeconomic factors and mental health among Vietnamese residents in Japan may also aid in the modification and development of health care policies. Therefore, to identify factors that may influence mental health status, the present study aimed to investigate symptoms of depression and anxiety among Vietnamese immigrants in Japan during the COVID-19 pandemic.

## Methods

This study was approved by the Kobe City College of Nursing Ethics Research Committee (approval number: 20124-05).

### Design and sampling

This cross-sectional survey was conducted among Vietnamese immigrants in Japan from September 21 to October 21, 2021. During the survey, Japan was in the latter half of the fifth COVID-19 wave. To prevent COVID-19 spread through contact, we conducted an online survey using SurveyMonkey. Since more than 70% of Vietnamese people use the Internet, [14] we conducted the survey using social networks as the main distribution platform. A one-page participants information statement, which also served as a recruitment poster, was posted in Facebook groups and on the personal accounts of the researchers. This statement briefly explained the study background, purpose, and procedure; voluntary nature of participation; anonymity of the questionnaire; strict privacy protection practices; and how to complete the questionnaire. It also included the statement, “By submitting the online questionnaire, the participant agrees that they have read and understand the participants information statement and agree to participate in the survey.” The inclusion criteria for participation were as follows: Vietnamese citizenship or Japanese citizenship in those of Vietnamese descent, current residence in Japan, and age ≥ 18 years. Participants received an online token for 200 Japanese yen ($1.8) after participation. To avoid duplicate responses from participants, SurveyMonkey was used to allow only one response from the same terminal.

### Data collection tools

The survey involved a self-administered questionnaire, which was used to collect data regarding demographic variables such as sex, age, duration of residence in Japan, area of residence, marital status, education level, birth country, visa status, Japanese language level, and pre-existing health conditions. Based on the opinions of Japanese infectious disease experts, we also classified residence into three categories based on the number of COVID-19 infections as of December 2021 (>2,000 per 100,000: Tokyo, Osaka, Okinawa; 1,999–1,000 per 100,000: Hokkaido, Saitama, Chiba, Kanagawa, Aichi, Kyoto, Nara, Hyogo, Fukuoka; <999 per 100,000: Others).

Economic and employment status was assessed based on national health insurance, public assistance, subjective socioeconomic status, and monthly income compared with those before the COVID-19 pandemic. International students were included in this survey. Under Japanese law, international students are allowed to work part-time or up to 28 hours per week outside their visa status. This income may account for a large portion of their living expenses. Therefore, in this survey, we also ascertained the income and working conditions of international students. Social support status was assessed based on whether patients had partners with whom they can talk about their health. Levels of depression and anxiety were assessed using the Patient Health Questionnaire-9 (PHQ-9) and Generalized Anxiety Disorder 7-item (GAD-7), respectively, which have been validated for assessing mental health status in Vietnam. [15–20] In our study, the Cronbach’s alpha values for the reliability of the PHQ-9 and GAD-7 were 0.844 and 0.933, respectively.

### Statistical analysis

We performed descriptive analyses using means and standard deviations (SD) for continuous variables as well as counts and percentages for binary or categorical variables. Each PHQ-9 item was rated on a four-point Likert scale ranging from 0 (not at all) to 3 (almost every day), with the total score ranging from 0 to 27. The severity of depression was divided into five categories based on PHQ-9 scores, as follows: none (0–4 points), mild (5–9 points), moderate (10–14 points), severe (15–19 points), and extremely severe (20–27 points). [21] A PHQ-9 score ≥10 points was considered to indicate clinically relevant depressive symptoms, based on a previous survey of COVID-19 in Vietnam. [15, 17–19] The severity of anxiety was classified into four categories based on GAD-7 scores: no anxiety (0–4 points), mild (5–9 points), moderate (10–14 points), and severe (15–21 points). [22] Each question item ranges from 0 (not at all) to 3 (almost every day) on a 4-point Likert scale, and the total score ranged from 0 to 21. GAD-7 scores ≥5 were considered to indicate clinically relevant symptoms of anxiety, as previously reported. [16, 20]

We conducted multivariable logistic regression analysis (forced-entry method) to identify factors related to symptoms of depression and anxiety, which were used as the dependent variables. The potential predictors included sociodemographic variables such as age, sex, marital status, education level, pre-existing health conditions; immigrant-specific variables such as the duration of residence in Japan, visa status, Japanese language proficiency; economic variables; variables related to the COVID-19 pandemic such as the rate of COVID-19 infection in the area; and variables related to social connectedness, such as the availability of a conversation partner. As employment status affects economic status, economic status, not employment status, was used as a predictor in this analysis. Inclusion of variables was based on previous studies of mental health among immigrants, [23, 24] and the chosen variables were entered simultaneously for multivariable regression. We created a correlation matrix before entering the independent variables; further, we confirmed that there were no strong correlations among the independent variables with r>0.80. Statistical analyses were performed using the SPSS software (version 19.0; IBM Corp., Armonk, NY, USA). Statistical significance was set at P<0.05 (2-sided tests).

## Results

A total of 1,046 people participated in the online survey, with 652 participants providing complete responses. After deleting responses with missing values or outliers, we included 621 responses. All participants were Vietnamese. The mean age (±SD) was 26.0±4.8 years; further, the mean duration of residence in Japan (± SD) was 3.4 ± 3.1 years. Approximately 30% of the respondents lived in areas with high infection rates. Most respondents (79.2%) were single (including divorced or bereaved), and almost all were born in Vietnam (99.0%). The most common visa status was “technical intern training (Ginojisshu)” (29.5%), followed by “international student” (29.3%) and “status of residence based on employment” (26.9%). The most common educational level was high school (38.3%), followed by college or university (34.5%) and technical school (21.3%). Further, 21.3% of respondents had a pre-existing health conditions, and the most common was gastrological (stomach/oesophagus/duodenum) conditions (5.5%), followed by otorhinolaryngological (5.2%) and musculoskeletal/joint disease (5.2%) conditions (Table 1).

**Table 1:**
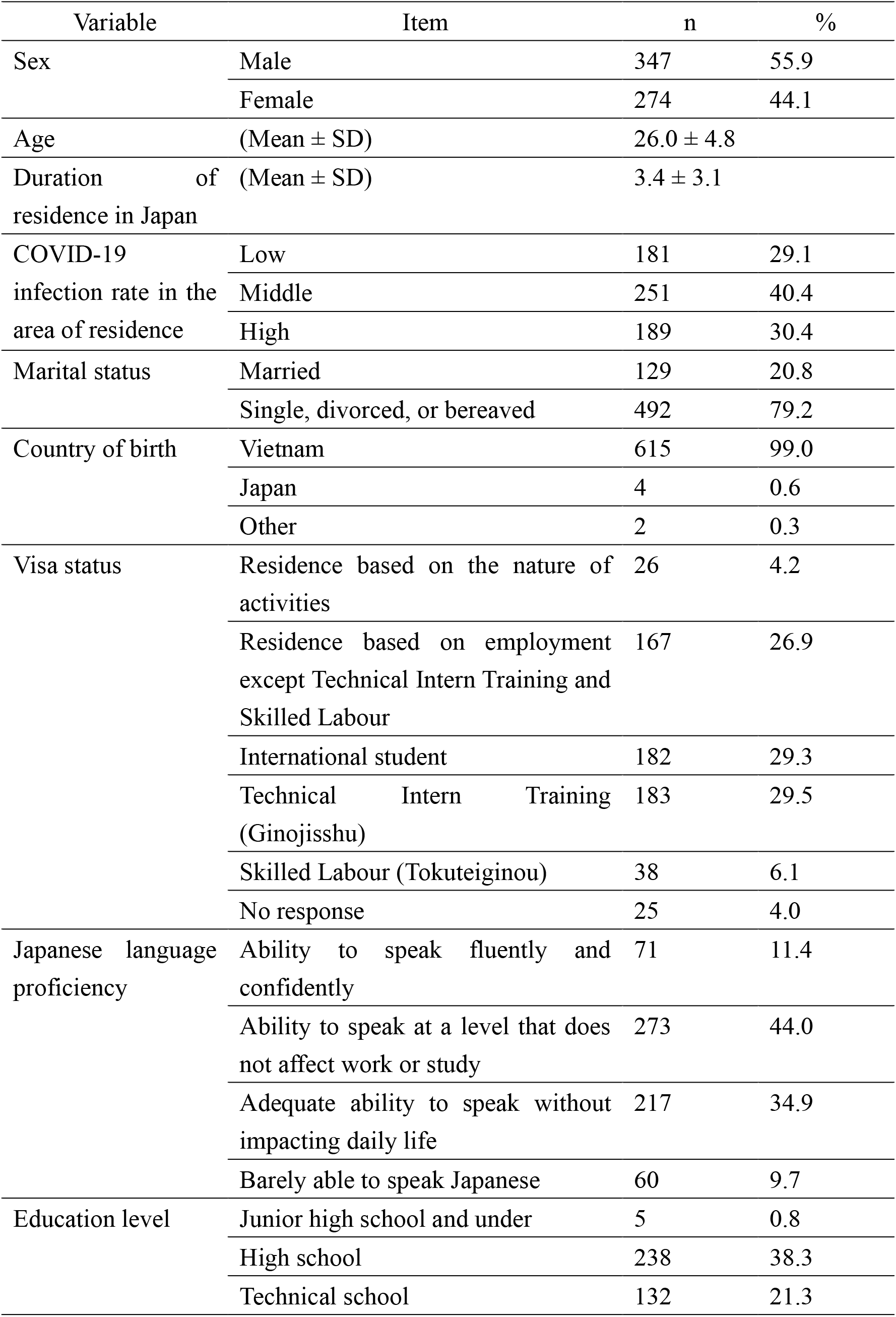
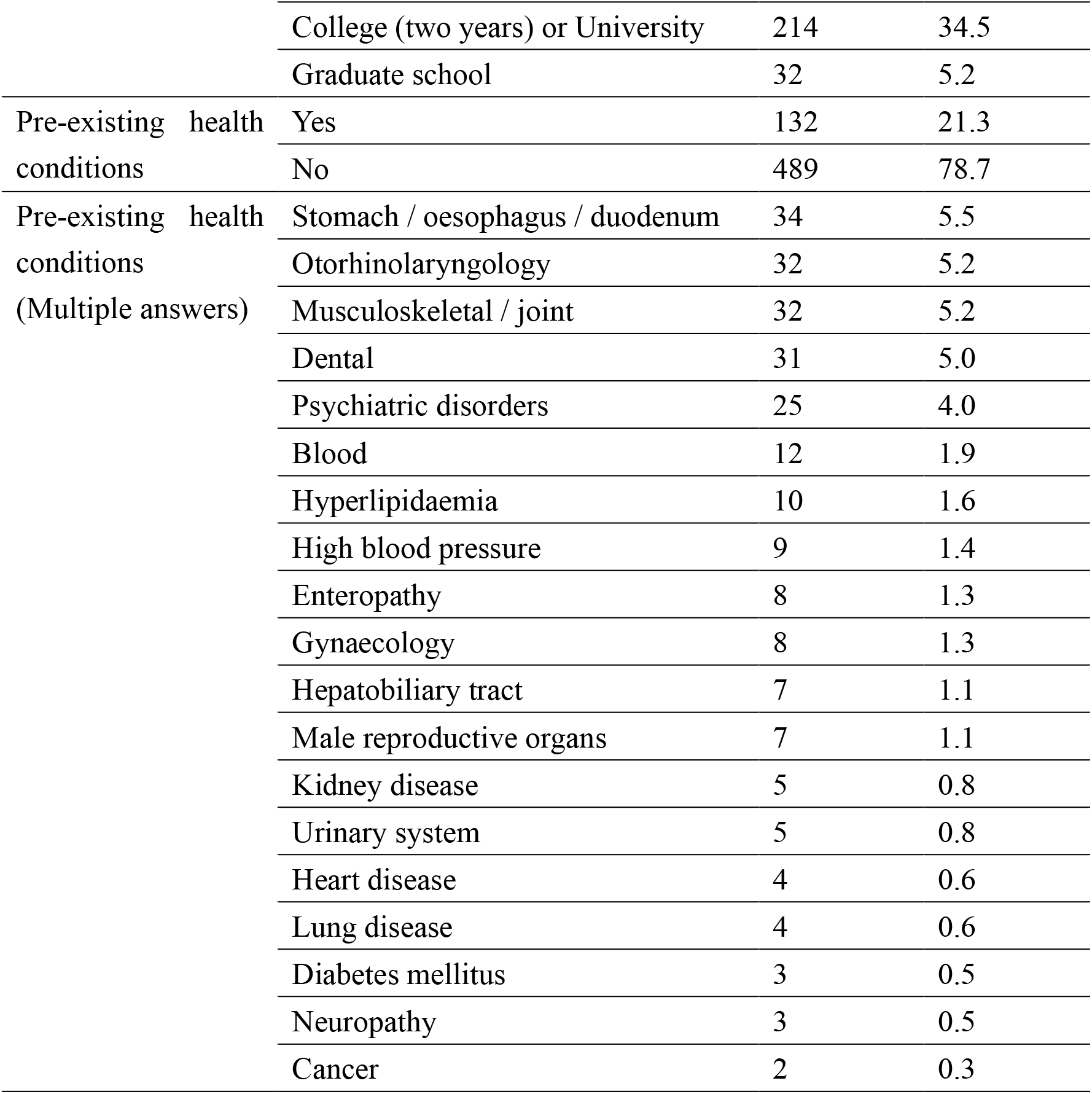
Participant characteristics (n = 621)

Regarding economic status, <5.5% and 6.9% of respondents lacked national health insurance and used public assistance, respectively. Approximately 34.6% of respondents reported a decrease in monthly income when compared with the period before the COVID-19 pandemic, with approximately 39.1% reporting a slight decrease. Approximately 14.2% and 46.2% of participants reported very poor and poor subjective socioeconomic status, respectively. Additionally, 18.7% of respondents reported dismissal from work/unemployment during the COVID-19 pandemic, while 64.1% reported a reduction in the number of working days (Table 2).

**Table 2:** Economic and employment status* (n = 621)

| Variable | Item | n | % |
| --- | --- | --- | --- |
| National health insurance | Yes | 587 | 94.5 |
|  | No | 34 | 5.5 |
| Public assistance | Used | 43 | 6.9 |
|  | Not used | 578 | 93.1 |
| Monthly income when compared with that before COVID-19 | Decrease of $\geq 40\%$ | 215 | 34.6 |
|  | Decrease of 10–40% | 243 | 39.1 |
| | Decrease $< 10\%$ | 146 | 23.5 |
| | Increase $> 10\text{--}40\%$ | 15 | 2.4 |
| | Increase $> 40\%$ | 2 | 0.3 |
| Subjective socioeconomic status | Really good | 5 | 0.8 |
|  | Slightly better | 47 | 7.6 |
|  | General | 194 | 31.2 |
|  | Slightly difficult | 287 | 46.2 |
|  | Very difficult | 88 | 14.2 |
| Employment status (multiple answers) | No change/no impact | 93 | 15.0 |
|  | Dismissal/Unemployment | 116 | 18.7 |
|  | Change in occupation type | 76 | 12.2 |
|  | Change in work content | 101 | 16.3 |
|  | Reduction in working days | 398 | 64.1 |
|  | Increase in working days | 17 | 2.7 |
|  | Other | 44 | 7.1 |

\* International students were included in the statistical analysis for “Economic and employment status” since they are allowed to have part-time jobs in Japan.

Further, 30.3% of participants reported having a partner with whom to discuss their health, with family (21.9%) being the most common, followed by Vietnamese friends (9.4%), work colleagues (8.7%), and medical professionals (including through online consultation) (4.3%) (Table 3).

**Table 3:** Social support status (n = 621)

| Variable | Item | n | % |
| --- | --- | --- | --- |
| Partner with whom to discuss health status | Yes | 188 | 30.3 |
|  | No | 433 | 69.7 |
| Who is the partner with whom you can discuss your health? (Multiple answers) | Family | 136 | 21.9 |
|  | Japanese friend | 21 | 3.4 |
|  | Vietnamese friend | 59 | 9.5 |
|  | People at the same office | 54 | 8.7 |
|  | People at the same school | 15 | 2.4 |
|  | Medical professionals (including through online consultation) | 27 | 4.3 |
|  | Religious facilities (temples, churches, etc.) | 4 | 0.6 |
|  | Other | 23 | 3.7 |

The mean PHQ-9 score was 7.9 ± 6.0. Mild, moderate, severe, and very severe depressive symptoms were observed in 212 (34.1%), 117 (18.8%), 52 (8.4%), and 34 (5.5%) participants, respectively. Further, 203 (32.7%) individuals had a PHQ-9 score ≥10 points. The mean GAD-7 score was 5.4±5.3. Mild, moderate, and severe symptoms of anxiety were observed in 169 (27.2%), 69 (11.1%), and 47 (7.6%) participants, respectively. Moreover, 285 (45.9%) individuals had a GAD-7 score ≥5 points (Table 4).

**Table 4:** Symptoms of depression and anxiety (n = 621)

| Variable | Item | n | % |
| --- | --- | --- | --- |
| Depressive symptoms (PHQ-9) | Mean ± SD | 7.9 ± 6.0 | |
|  | No depression (0–4 points) | 206 | 33.2 |
|  | Mild (5–9 points) | 212 | 34.1 |
|  | Moderate (10–14 points) | 117 | 18.8 |
|  | Severe (15–19 points) | 52 | 8.4 |
|  | Very severe (20–27 points) | 34 | 5.5 |
|  | 10–27 points | 203 | 32.7 |
| Anxiety symptoms (GAD-7) | Mean ± SD | 5.4 ± 5.3 | |
|  | No anxiety (0–4 points) | 336 | 54.1 |
|  | Mild (5–9 points) | 169 | 27.2 |
|  | Moderate (10–14 points) | 69 | 11.1 |
|  | Severe (15–21 points) | 47 | 7.6 |
|  | 5–21 points | 285 | 45.9 |

We performed a multivariable logistic regression analysis by the forced-entry method. Twenty-five participants with missing responses for residency status were considered as missing values. The results of the χ^2^ test were significant (P<0.001) for the PHQ-9 and GAD-7 regression models. The Hosmer and Lemeshow’s test for PHQ-9 and GAD-7 scores revealed P values of 0.671 and 0.194, respectively, and the goodness-of-fit for the model was acceptable. Additionally, there were no outliers in which the predicted values exceeded the measured values by ±3 SD. Depressive symptoms were associated with age (odds ratio [OR]=0.94, 95% confidence interval [CI]=0.89–0.99), pre-existing health conditions (OR=2.46, 95% CI=1.61−3.76), and low socioeconomic status (OR=2.47, 95% CI=1.64–3.71). Factors related to anxiety included being single (OR=1.72, 95% CI=1.01–2.93), pre-existing health conditions (OR=2.52, 95% CI=1.63–3.88), low socioeconomic status (OR=2.72, 95% CI=1.87–3.97), and not having a partner with whom to discuss one’s health (OR=1.66, 95% CI=1.11–2.47) (Table 5).

**Table 5:**
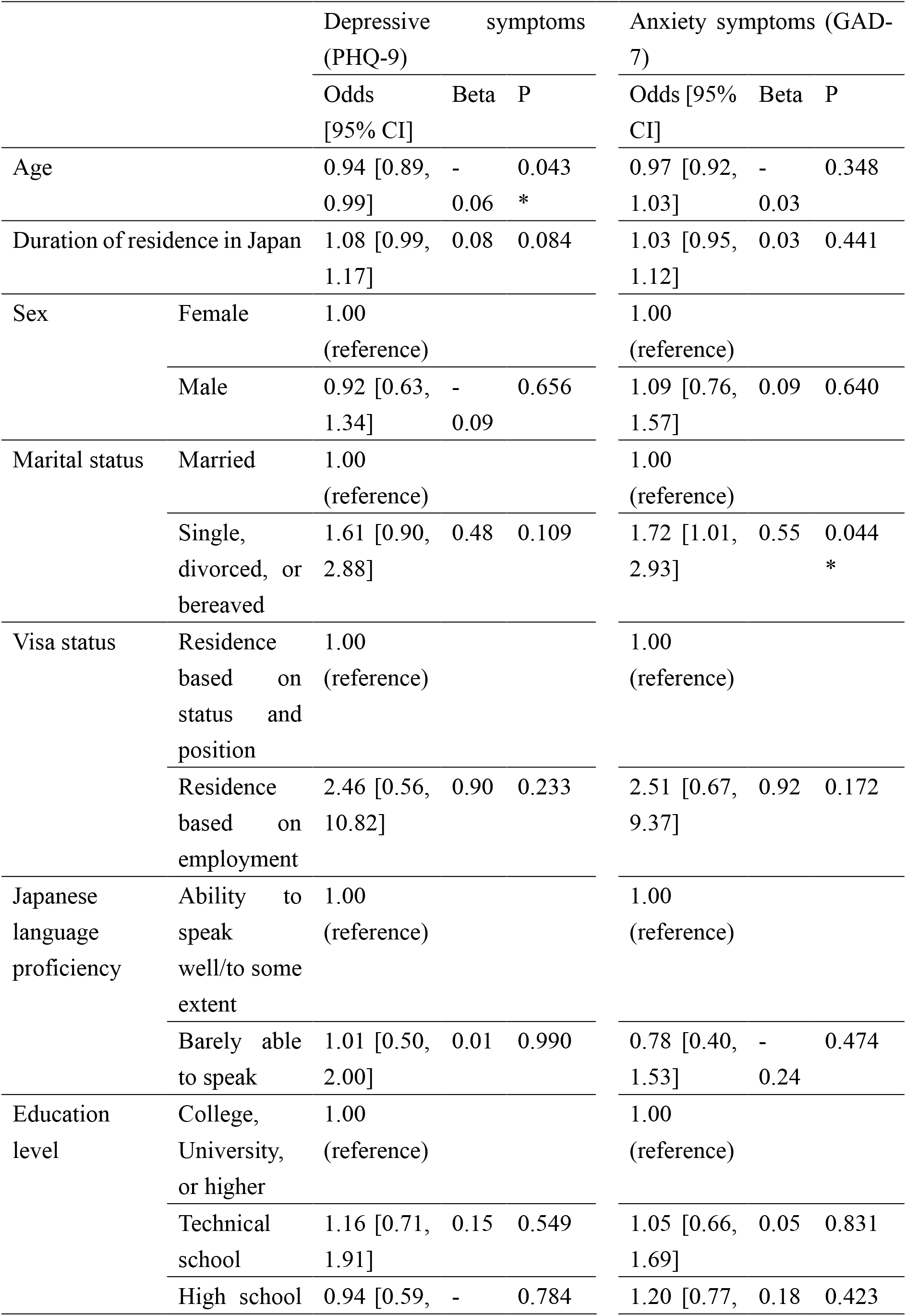
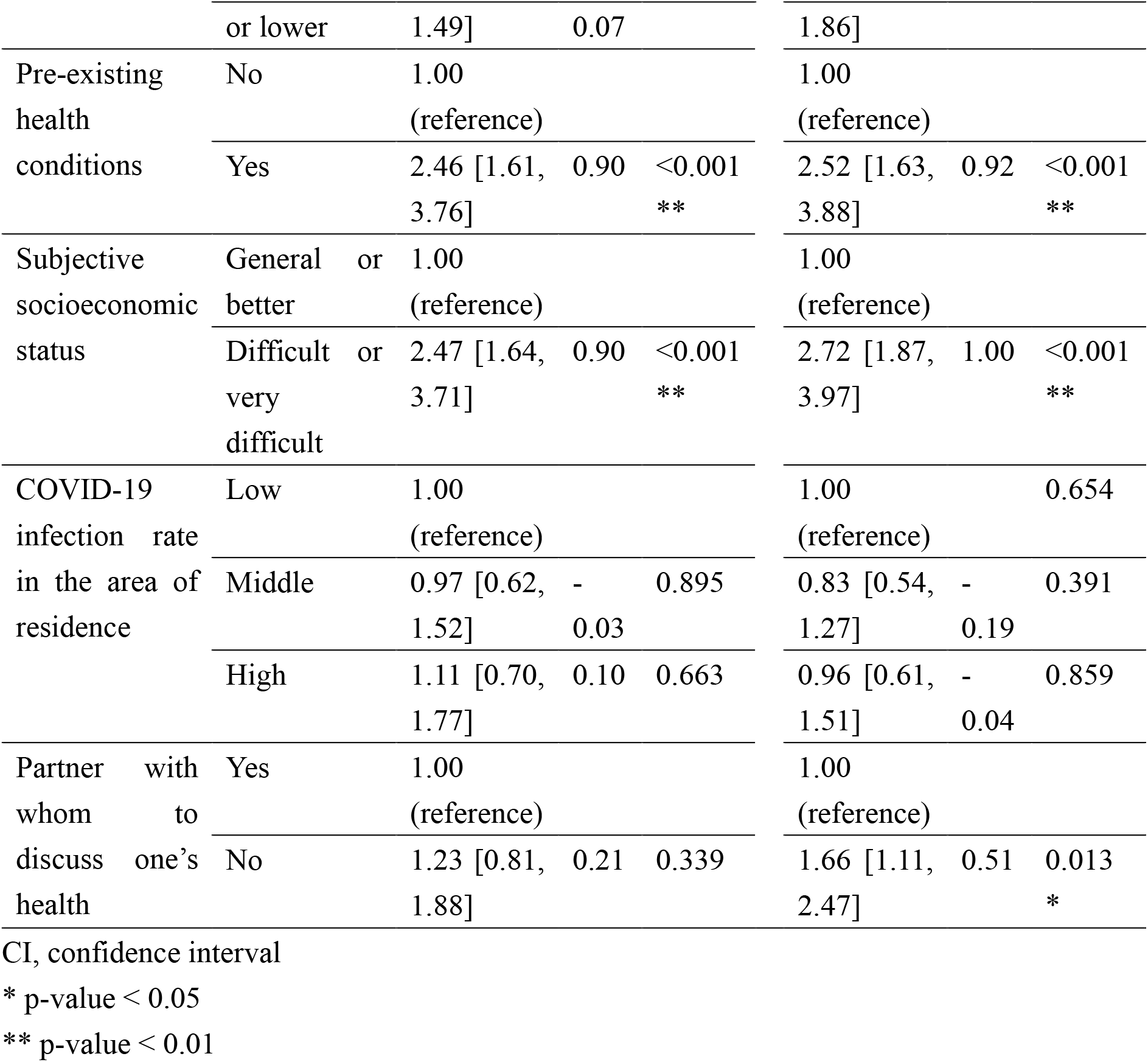
Factors associated with symptoms of depression and anxiety (n = 596)

## Discussion

Our findings indicated that many Vietnamese immigrants in Japan experienced a decrease in income and worsening working conditions during the COVID-19 pandemic. In addition, approximately half of respondents reported symptoms of depression and anxiety, which were significantly associated with pre-existing health conditions, and low socioeconomic status in the multivariable regression analysis. Beyond these associations, symptoms of depression were associated with age, and the symptoms of anxiety were associated with the availability of partners with whom to discuss one’s health. These results demonstrate that there is a mental health problem among Vietnamese immigrants in Japan, highlighting the need for financial support and measures for preventing social isolation.

We observed a high prevalence of depression symptoms, with 61.3% and 32.7% of respondents reporting mild-to-severe and moderate-to-very severe symptoms, respectively. This prevalence is higher than previously reported values. The prevalence of depressive symptoms was greater in our study than in other studies conducted after the start of the pandemic. Porter et al. [25] reported that the prevalence of mild-to-severe depression as measured using the PHQ-8 was 9.5% among the young population, while Luong et al. [26] reported a prevalence rate of 18.4% for moderate-to-severe depression (PHQ-9 scores) among pregnant women in Vietnam. Other studies using the PHQ-9 have reported that the rate of moderate-to-severe depressive symptoms was higher among Vietnamese immigrants than among Japanese citizens: 17.9% among adults aged 18 years and older in 2020; [27] 11.6% in 2020 and 16.6% in 2021 among college students; [28, 29] and 14.8% among hospital workers in 2020. [30] The current evidence indicates that depressive symptoms among Vietnamese immigrants in Japan appear to be more severe than those among Japanese and Vietnamese individuals living in their home countries.

We observed a high prevalence of anxiety symptoms in the current study, with 45.9% and 18.7% of respondents reporting mild-to-severe and moderate-to-severe anxiety symptoms, respectively. A previous study reported that only 33.1% and 5.5% of clinical health care workers in Vietnam experienced mild-to-severe and moderate-to-severe symptoms of anxiety, respectively. [20] Another recent study noted that 25.5% and 4.3% of healthcare workers in Vietnam exhibited mild-to-severe and moderate-to-severe anxiety symptoms, respectively. [16] Ueda et al. [31] reported mild-to-severe symptoms of anxiety in 31.6% of Japanese people 18 years and older, based on GAD-7 scores. As observed for depressive symptoms, the prevalence of anxiety appears to be similar between Vietnamese immigrants and other immigrant populations in Japan, with immigrants experiencing more severe symptoms than Japanese and Vietnamese individuals living in their home countries.

The World Health Organization recently reported that economic problems have affected the mental health of migrants during the COVID-19 pandemic. [32] In December 2020, approximately 46,000 foreigners living in Japan were laid off, unable to continue working, or unable to return home, [4] many of whom were likely Vietnamese. In this survey, 60.4% of participants reported a slightly difficult or worse economic situation relative to the pre-pandemic period, while 14.2% reported a severe change in economic status, indicating that Vietnamese immigrants in Japan remain anxious about their current and future lives in Japan. The Ministry of Health, Labour, and Welfare (Japan) has been providing financial support including a welfare fund loan system, public assistance, counselling, support for finding employment, and support for early return to the home country; [4, 33, 34] however, there have been no positive outcomes. Nonetheless, there is a need to continue supporting the economic and daily life of Vietnamese immigrants in Japan since worsening economic situations and instability in daily life are strongly associated with mental health status.

Our findings additionally demonstrated that the presence of pre-existing health conditions significantly impacted mental health among Vietnamese immigrants in Japan. Among the most important health issues in Japan is the health disparity among immigrants living in Japan given the lack of access to medical services. [35] Another report on immigrants in Japan who participated in free medical check-ups conducted by a Non-Governmental Organization revealed that many were not connected to medical services. [36] Fear of COVID-19 has also generally reduced the number of people receiving medical services in Japan. [37] The COVID-19 pandemic further restricted access to medical services among immigrants in Japan, including Vietnamese residents, which might contribute to increases in their anxiety.

Our analysis also revealed that age differences have a significant impact on depressive symptoms. The result was that depressive symptoms were less likely to occur with increasing age. This means that young people are at high risk. Similar reports have been seen in Vietnam and Japan under the COVID-19 pandemic; [25, 38] therefore, this result may not be a characteristic issue for Vietnamese immigrants in Japan. However, young Vietnamese individuals in Japan find it difficult to respond to changes in their living environment due to COVID-19 pandemic, which is expected to lead to an increase in their depressive symptoms. Our analysis also revealed that being single and absence of partners with whom to discuss one’s health significantly affected the risk of experiencing symptoms of anxiety. The Immigration Services Agency of Japan reported that foreigners living in Japan had few connections with others outside of work. [39] A South Korean study conducted in October 2020 reported that immigrants who live alone have moderate-to-extreme anxiety symptoms when compared with those living with their families. [11] These findings indicate that connections with others, especially family members, are important for maintaining mental health among immigrants. Thus, Vietnamese immigrants in Japan will require further support to connect with their families, Vietnamese friends, and others during the remainder of the COVID-19 pandemic.

The present study had some limitations, including its self-reported and retrospective nature, which may have introduced recall bias. In addition, the survey may not be fully representative of the Vietnamese population living in Japan since some older adults of Vietnamese people may not have been able to participate using the online format. Since this survey was collected via social media, responses may have been biased toward Vietnamese residents in Japan who are interested in mental health. Therefore, there is a possibility of participant selection bias. Finally, the study’s research design did not consider the association between family networks and mental health at the time of immigration noted by Dykxhoorn et al. [40] Further research is needed to clarify the actual mental health status of Vietnamese people living in Japan by re-examining the selection of participants and confounding factors.

## Conclusions

In this cross-sectional survey, we examined the mental health status of 621 Vietnamese immigrants living in Japan during the COVID-19 pandemic. Our analysis yielded the following five main findings: (1) many Vietnamese immigrants experienced economic difficulties; (2) symptoms of depression (moderate-to-very-severe) and anxiety (mild-to-severe) were observed in 32.7% and 45.9% of respondents; (3) low subjective socioeconomic status and pre-existing health conditions were associated with depression and anxiety; (4) younger people are more likely to have depressive symptoms; and (5) being single and the absence of partners with whom to discuss one’s health were associated with anxiety. Together, our findings demonstrated that mental health status was worse among Vietnamese immigrants than among Japanese citizens during the COVID-19 pandemic, and that there is a need for financial support and measures that can prevent poor mental health status among them.

## Data Availability

The data that support the findings of this study are available from the corresponding author upon reasonable request.

## List of Abbreviations

CI: confidence interval
GAD-7: Generalized Anxiety Disorder-7
OR: odds ratio
PHQ-9: Patient Health Questionniare-9
SD: standard deviation

## Acknowledgments

We wish to extend special thanks to all the respondents who participated in the survey. We also thank Professor Hiroya Matsuo for his helpful advice.

## Author contributions

T. Y. conceived the original idea for the study. T. Y., P. N. Q., S. I., and K. K. designed the questionnaire. T. Y., C. Y., and K. K. analysed the data and wrote the first draft of the manuscript. P. N. Q., N. E., E. S., C. Y., S. I., K. S., and K. K. contributed to the interpretation of the results. All authors made substantial intellectual contributions to the study and approved the final draft of the manuscript.

## Consent for publication

Not applicable.

